# The illusion of personal health decisions for infectious disease management: disease spread in social contact networks

**DOI:** 10.1101/2023.01.01.22284075

**Authors:** Matthew Michalska-Smith, Eva A Enns, Lauren A White, Marie L J Gilbertson, Meggan E Craft

**Affiliations:** Department of Ecology, Evolution and behaviour, University of Minnesota, USA; Department of Plant Pathology, University of Minnesota, USA; School of Public Health, University of Minnesota, USA; National Socio-Environmental Synthesis Center, University of Maryland, USA; Veterinary Population Medicine Department, University of Minnesota, USA

## Abstract

Close contacts between individuals provide opportunities for the transmission of diseases, including COVID-19. Individuals take part in many different types of interactions, including those with classmates, co-workers, and household members; the conglomeration of all of these interactions produces a complex social contact network interconnecting individuals across the population. Thus, while an individual might decide their own risk tolerance in response to a threat of infection, the consequences of such decisions are rarely so confined, propagating far beyond any one person. We asses the effect of different population-level risk-tolerance regimes, population structure in the form of age and household-size distributions, and different types of interactions on epidemic spread in plausible human contact networks to gain insight into how contact network structure affects pathogen spread through a population. In particular, we find that changes in behaviour of vulnerable individuals in isolation is insufficient to reduce those individuals’ infection risk, that population structure can have varied and counter-acting effects on epidemic outcomes, and that, in general, interactions among co-workers have a greater contribution to disease spread than do interactions among children at school. Taken together, these results promote a nuanced understanding of disease spread on contact networks, with implications for public health strategies.

## 1 Introduction

Many respiratory diseases, including influenza, tuberculosis, and COVID-19, are primarily transmitted through close contact between an infectious individual and a susceptible one, whether by direct physical contact or through expelling contaminated droplets via coughing, sneezing, or breathing [1]. While not all such interactions lead to a transmission event, the transmission network (*i.e*. the actual set of who infects whom in a population) is a subset of this wider contact network (*i.e*. the set of all interactions between individuals that could result in in disease transmission) [2].

The importance of interpersonal contact for disease dynamics has been recognized for centuries, with isolation of infected individuals being recorded in fifteenth century Italy [3], and has become more formalized in recent decades [4, 5, 6]. Yet, detailing the specific ways in which the structure of contact networks relates to differences in disease spread between populations has been hampered by the size and complexity of human social networks, which are an agglomeration of many different kinds of interpersonal interactions [7]. A given person, for instance, will interact with some people at home (their family or housemates), others when they go to work (co-workers and colleagues), and yet others when they go to the local store for groceries (neighbors and strangers). Not only do the individuals involved in each of these sub-networks differ for any given person, but also the structure and intensity of interactions might likewise differ between contexts.

Pathogens spread differently in different localities in part because of a difference in social contact network structure [8, 9, 10, 6], thus we might also expect disease dynamics to vary across social contexts: to spread differently at work than at school, through a home than through a neighbourhood. Yet, unlike the case of two distinct localities, these layers of interactions are also not independent from one another, linked by the individuals that take part in multiple layers. It is the combination of these layers into an integrated network detailing all possible infection pathways that affects the ultimate spread of disease through a population. But how much does each type of interaction contribute to this final disease spread? Can the layers be modified independently in order to alter a population’s risk in the face of disease spread?

Operationalizing the connection between contact network structure and disease spread, public health interventions such as travel restrictions, business and school closures, and individual isolation and/or quarantining seek to reduce disease spread through direct modification of the contact network [11, 12]. In short, such modifications seek to sever potential infection pathways through the contact network before they are realized, limiting the number of potential secondary cases available to a given infectious individual. These approaches can range from hyper-local—only isolating individuals who have been confirmed to be infected—to society-wide—wholesale economic lockdowns and *cordons sanitaire* [13].

In their initial response to the COVID-19 pandemic, many countries imposed strict restrictions on social interactions—especially those within schools and workplaces [14]—with the goal of limiting disease spread through the mass fragmentation of societal contact networks [15]. While such efforts have, in general, been found to be effective both historically [16], and in the current pandemic [17, 18], they are nevertheless a blunt intervention. More restrained approaches, such as test-trace-quarantine can be more surgical in their application, but their efficacy tends to be limited by insufficient participation and high costs when cases are surging [4, 19, 20]. A middle ground could involve restricting certain types of interactions while leaving others unaffected, balancing disease mitigation and socio-economic hardship (*e.g*. closing schools, but leaving workplaces open, or *vice versa*). Finally, not all public health interventions seek to completely sever links in the contact network. Softer approaches, such as masking, increased attention to personal hygiene, improved ventilation, and physical distancing can be used to reduce the strength of interactions, *i.e*. reduce the transmission rate given interaction between two individuals, rather than eliminating the interaction altogether [21, 22].

In addition to differences between types of interactions, which might be relatively consistent from one individual to another, there are also differences between individuals both in behaviour [23, 24] and in underlying health conditions that increase the likelihood of experiencing adverse health outcomes in the event of infection [25]. While a decision might be made on a personal level (*e.g*. one person might decide to return to in-person work, while another might take advantage of a work-from-home option), the consequences of this decision have the potential to propagate far beyond a focal individual, with individuals serving as either bridge or firewall in a pathogen’s infection chain.

In this work, we investigate the impact of plausible human contact network structure [26, 7, 13] on the spread of disease across three scales of network structure, using COVID-19 as an example. First, we consider differences in individual risk tolerance with respect to an individual’s contact with persons in the network who are at greater risk of adverse outcomes following infection (*i.e*. “vulnerable” individuals). Second, we consider the effect of wider population structure on the spread of disease, comparing two locales that differ in age- and household-size distributions. Finally, we add to these two considerations the relative contribution of two layers in the contact network (*i.e*. interactions between classmates at school and interactions between co-workers at work). We focus on these two layers in particular as they (along with household interactions) comprise the majority of potential transmission events in modern society [27], and have been the focus of prior research and public health interventions, better allowing us to contextualize any results [20, 14, 13, 28]. Taken together, the results of this investigation provide a foundation for better understanding the role of contact network structure on the spread of disease, and an avenue for better targeting public-health interventions to limit further disease spread.

## 2 Methods

### 2.1 Network construction

We constructed human contact networks by sequentially adding interaction layers to a base network of individuals grouped into households according to United States (US) 2019 American Community Survey data on the distribution of household sizes [29]. Each individual was assigned an age (according to US 2019 American Community Survey data [30]) and a binary “vulnerable” status. Vulnerability was assigned according to age-adjusted hospitalization rates [31]. School-age children were then assigned to classrooms (using an approximate classroom size of 20 students), and pre-retirement-age adults (accounting for US unemployment rates) to workplaces (according to a modified distribution of US business sizes). To make our networks more realistic, we additionally considered the effect of community spread of disease outside of the structured settings of work and school (*e.g*. spread at the grocery store or local shopping center). For this, we added a layer connecting all individuals in the network to all others at a low transmission rate (*i.e*. “background transmission”).

Each of these four network layers is a collection of distinct, fully connected sub-networks that correspond to households, classrooms, workplaces, or the community as a whole. By layering these networks together, the isolated clusters from any one layer become intertwined through the connections in other layers. For example, a student might be connected to an unrelated, vulnerable adult through an interaction chain involving a classmate interaction with a friend, a household interaction between the friend and their parent, and a workplace interaction between the parent and an elderly co-worker. The strength of interaction in the co-worker and classmate interaction layers was varied systematically to explore the relative importance of each of these layers, while those in the household layer (as well as background transmission) were held constant.

We considered two US states as case studies for comparing differences in local population structure. Using US 2019 American Community Survey data (see Supplementary Information section S1 for detailed data sources), we constructed synthetic networks with age- and household-size distributions matching those of either Florida—a US state with a relatively high average age and small average household size—or Texas—a US state with a relatively low average age and large average household size (section S2 and fig. S1). Each network was further populated with classmate and co-worker interaction layers, as detailed above, using the same algorithm and parameters for both localities. Networks were generated to have approximately the same number of individuals (3 000), which necessitates a different number of households in each network due to the aforementioned differences in average household size.

Finally, we modified the above networks according to three risk-tolerance scenarios, generalizing behaviours to all individuals in the population. First, we considered a population in which all individuals, regardless of inherent vulnerability, behave identically, fully participating in their co-worker and classmate interactions. Second, we considered a case where vulnerable individuals avoid those interactions (*i.e*. do not go to work/school and therefore have no co-worker or classmate interactions) in order to reduce their own exposure risk. Finally, we considered a case where all members of any household containing at least one vulnerable individual avoid co-worker and classmate interactions in an effort to reduce exposure to their vulnerable housemates. Table 1 details differences between the networks constructed for each of the two locales and under different risk-tolerance scenarios.

**Table 1:** Summary statistics for networks generated for each of the two localities used in the main text.

| Metric | “Florida” mean (sd)<sup>1</sup> | “Texas” mean (sd)<sup>1</sup> |
| --- | --- | --- |
| Number of individuals | 3 001 | 3 000 |
| Number school-age | 503 (20.5) | 648 (22.5) |
| Number employed | 1 549 (27.4) | 1 595 (27.5) |
| Number vulnerable | 628 (22.2) | 535 (20.9) |
| Number of households | 1 212 | 1 056 |
| Number households with children | 462 (15.1) | 517 (13.4) |
| Number of households with vulnerable | 508 (16.3) | 423 (14.9) |
| Total number of edges (no contact avoidance)<sup>2</sup> | 24 961 (611.1) | 28 411 (578.5) |
| Household edges | 3 425 | 4 519 |
| Classmate edges | 5 890 (307.7) | 7 744 (345.0) |
| co-worker edges | 15 646 (593.2) | 16 148 (560.5) |
| Edges when vulnerable individuals avoid work/school interactions | 17 655 (564.1) | 20 766 (570.6) |
| Edges when members of vulnerable households avoid interactions | 7 857 (539.2) | 8 605 (610.1) |

<sup>1</sup> Values are presented with both mean and standard deviation except when there was no variance, in which case the constant value is presented.
<sup>2</sup> “Background” transmission links are omitted from this (and other edge) count(s). Because they connect every individual to every other, there are always $N(N - 1)/2$ such edges, where $N$ is the number of individuals in the network.

### 2.2 Disease simulation

Pathogen spread through the population was simulated according to modified SEIR dynamics, using a discrete-time, chain binomial model [32]. Specifically, individuals (nodes) in the network fell into one of six classes at each timestep: susceptible to infection (*S*), exposed but not yet infectious (*E*), infectious and symptomatic (*I*_*s*_), infectious and asymptomatic (*I*_*a*_), recovered and immune to future infection (*R*), or a victim of disease-induced mortality (*D*). Transitions between classes were governed by rate parameters (table 2) that, when appropriate, could take two discrete values based on an individuals’ inherent vulnerability to severe disease.

**Table 2:** Estimates and description of parameters for the SARS-CoV-2 model used in this work.

| Parameter | Description | Point Estimate* | Reference(s) |
| --- | --- | --- | --- |
| $\beta_{background}$ | The transmission rate for community interactions | 0, 0.001/$N$, or 0.1/$N$ | |
| $\beta_{household}$ | The transmission rate for interactions between household members | 0.13 | Rosenberg et al. [35],<br>Bar-On et al. [36] |
| $\beta_{classmate}$ | The transmission rate for interactions between classmates | none<sup>†</sup> | |
| $\beta_{co-worker}$ | The transmission rate for interactions between co-workers | none<sup>†</sup> | |
| $\sigma$ | The incubation period (the inverse of the average latent period duration) | 1/5.5 | Bar-On et al. [36] |
| $\rho$ | Proportion of new infectious individuals that are symptomatic | 0.35<sup>‡</sup> | Wang et al. [37],<br>Bar-On et al. [36] |
| $\gamma$ | The recovery rate (the inverse of the average duration of the infectious period) | 1/4.5 | Bar-On et al. [36] |
| $\mu$ | Baseline mortality rate for symptomatic, non-vulnerable individuals | 1/27 000 | The World Bank [38],<br>Ugarte et al. [39] |
| $\nu$ | Additional mortality due to vulnerability | 1/1 000 | Bar-On et al. [36] |

\* Units are days<sup>-1</sup> unless otherwise noted; $N$ signifies the number of individuals in the network.
<sup>†</sup> values sampled from $10^{[-3, -0.5]}$ (one for each transmission rate per simulation), where $[a, b]$ represents a range from $a$ to $b$ (inclusive) from which values were uniformly randomly sampled.
<sup>‡</sup> this value is unit-less

Explicitly, susceptible nodes can be infected at each timestep depending on their network connections:

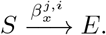

Where 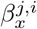 is the rate of transmission between two nodes, one infectious (*i*) and one susceptible (*j*), connected by interaction type *x*. A susceptible individual will have the chance to be infected by each of their infectious interaction partners on each timestep. We considered three alternative rates of background transmission (table 2), but only present figures corresponding to a value of *β*_*background*_ = 0.001*/N* (where *N* is the number of nodes in the network) in the main text. See Supplementary Information section S3 for figures corresponding to values of 0 and 0.1*/N*.

Exposed individuals’ experience disease progression at a constant rate:

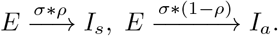

Where *σ* represents the disease progression rate (the inverse of the time between becoming infected and becoming infectious) and *ρ* is the proportion of infected individuals that develop symptoms. Infectious individuals recover or die at constant rates (depending on their symptomaticity and inherent vulnerability):

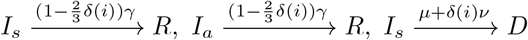

Where *δ*(*i*) is an indicator function that returns 1 if an individual is vulnerable, and 0 otherwise, *γ* represents the rate of recovery, which is (approximately three times) longer for vulnerable individuals [33, 34], *μ* represents a baseline mortality rate, and *ν* represents additional mortality experienced by vulnerable individuals. All disease parameters were set to literature values approximated for the initial wave (original Wuhan strain) of COVID-19 (table 2).

Note that we assume: 1) per-contact transmission rates are independent of the symptomaticity of the infectious interaction partner, 2) all mortality is disease induced, and 3) that only symptomatic individuals suffer disease-induced mortality.

Populations were seeded with a single infected individual and simulations were allowed to run until no further infections were possible. A total of 10 000 unique combinations of classmate (*β*_*classmate*_) and co-worker (*β*_*co−worker*_) transmission rates were sampled using a Latin-Hypercube approach [40], each parameter combination was run for each of the two localities and three risk-tolerance regimes, leading to a total of 60 000 simulated epidemics.

### 2.3 Epidemic outcome quantification

Epidemic spread was quantified using the total number of individuals infected, the total number of vulnerable individuals infected, the average number of individuals concurrently infectious, the total number of individuals that died, the maximum number of concurrently infectious individuals, the number of timesteps to reach that peak, and the number of timesteps that passed before the first vulnerable individual was infected.

All simulations were conducted in C++ version 8.1.0, with data manipulation and plotting done in R version 4.2.0 [41]. For specific packages used, see Supplementary Information section S4. Code to replicate all aspects of these analyses is available online: https://github.com/mjsmith037/Layered_Interactions_COVID_Model.

## 3 Results & discussion

### 3.1 Quantifying the effect of differential risk-tolerance behaviour

As expected, increasing the transmission rate for classmates or co-workers increased the number of infectious individuals, the number of vulnerable people infected and the total number of individuals that died (fig. 1). Yet, this effect was modulated by behaviour: in particular, we found that the actions of vulnerable individuals in isolation did little to reduce the total disease burden on the population in terms of number of cases and deaths, except when transmission rates were already low. However, there was a substantial reduction in these values when household members likewise avoided work/school interactions themselves from other individuals in the network (fig. 1). This trend was consistent across metrics of epidemic outcome, such as the peak infection prevalence, the number of vulnerable individuals infected, and the total number of deaths. Likewise, we saw consistency across a range of transmission rates between classmates and between co-workers, though if both rates were sufficiently low (bottom left corner of each panel in fig. 1), the extent of disease spread was minimal. Importantly, if background transmission rates are high enough, classmate and co-worker transmission is rendered irrelevant, precluding any differences between behaviour treatments (Supplementary Information fig. S5). We emphasize that these effects are not simply a result of reducing the number of interactions in the network. Repeating the above simulations (*i.e*. removing the same number of edges from each interaction layer in the network as above), but choosing which links within each layer to remove at random (*i.e*. irrespective of an individuals (contact with) vulnerable individuals), yields no qualitative difference between link-removal treatments (section S5 and fig. S8). Finally, these patterns were also consistent across local population structures (*i.e*. Texas or Florida), so we aggregated results across locales for figs. 1, S2, S5 and S8.

**Figure 1:**
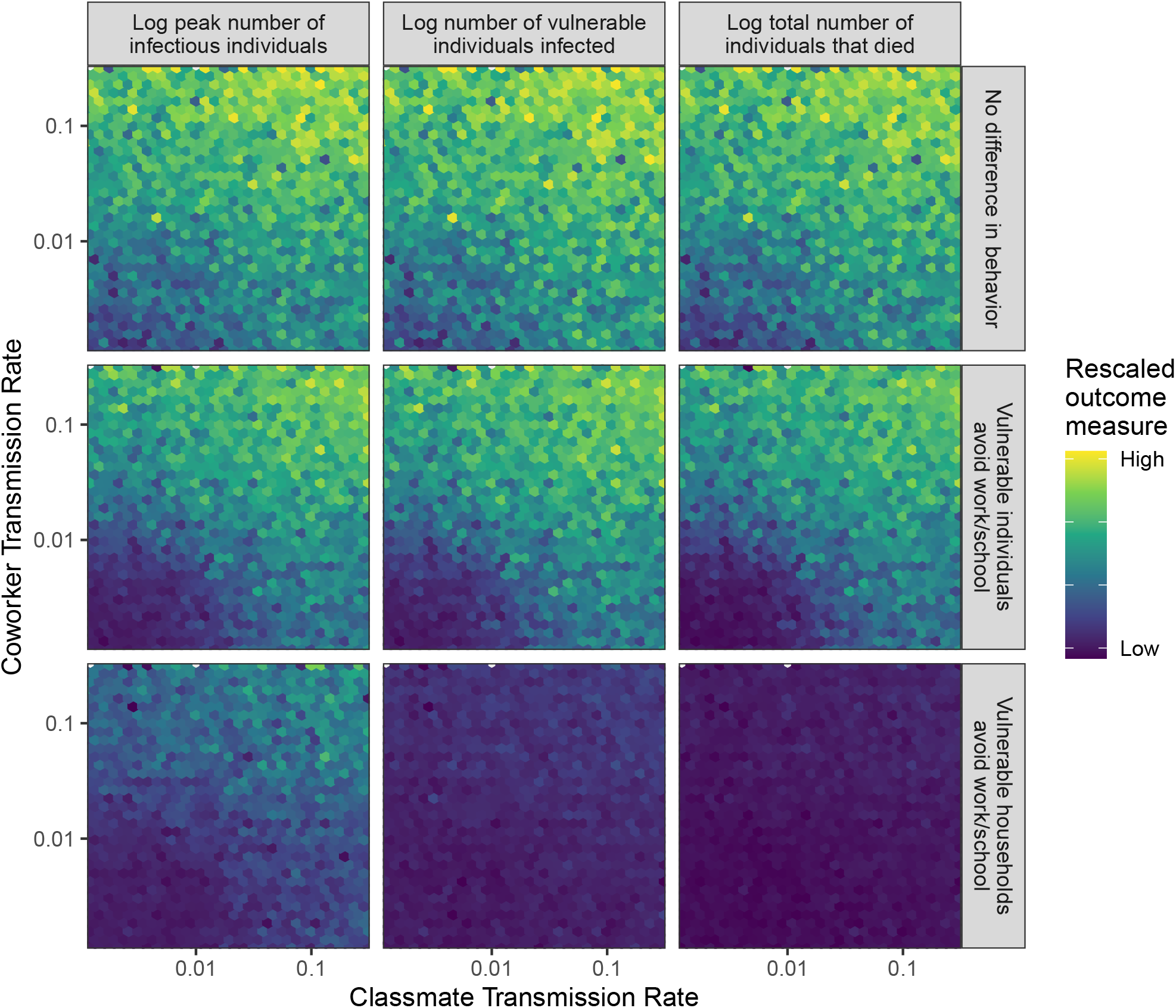
Relative epidemic outcome (columns), quantified as the peak number of individuals infected (left), the number of vulnerable individuals infected (middle), or the total number of individuals that died (right) over the course of the simulation. Individuals in the network either (rows): did not change behaviour in response to (contact with individuals with) vulnerability status (top), changed behaviour if they were vulnerable themselves (middle), or changed behaviour when a member of their household was vulnerable (bottom). Multiple points within each hexagon were averaged to produce the plotted value. Mean values were then log-scaled and normalized for each epidemic outcome such that the maximum value is 1 (yellow) and the minimum value is 0 (purple). Each panel consists of a heatmap showing the relative epidemic outcome of simulations spanning various levels of co-worker (vertical axis) and classmate (horizontal axis) transmission. Maximum disease burden in all cases occurs when the transmission rates are both high (top right corners of each panel), while the disease tends to die out with minimal cases and death when both rates are low (bottom left of each panel). Results here are aggregated across local population structures, which were qualitatively similar. See Supplementary Information (section S3 and figs. S2 and S5) for analogous figures under different background transmission rates.

The absence of reduced disease burden when only vulnerable individuals change their behaviour can be attributed, at least in part, to the high-interaction strength expected for within-household interactions, limiting the efficacy of contact-reduction for vulnerable individuals sharing households with less vulnerable individuals. Unless the whole household takes actions to reduce their exposure, we see limited benefits of reducing a particular individuals exposure in isolation. This is true even if we only look at the rates of infection in the vulnerable individuals themselves. Moreover, because the vast majority of deaths from COVID-19 are individuals with underlying health conditions that provide an inherent vulnerability to adverse outcomes [42], reducing the number of vulnerable individuals infected has a direct effect of reducing mortality as well.

It is important to also note that not all interaction decisions are the product of (or even align with) a particular individual’s risk tolerance, but rather are the combined product of individual decisions and systemic social and workplace structures that constrain individual behaviour. This is a critical consideration in the construction of policy, especially when such policies tend to be focused on individuals themselves and (occasionally) those directly under their care, rather than a consideration of potential interactions with (and consequent transmission risk to) other vulnerable individuals [43]. For instance, those with underlying health conditions might be able to apply for remote work with a note from a medical provider, however, they are less likely to be granted accommodation if their housemate is the vulnerable individual. Relatedly, such policies have historically been applicable only after an individual is infected, rather than allow for the reduction of transmission prophylactically. More effective protection of vulnerable individuals would require facilitating household-wide action to reduce exposures [44, 45].

### 3.2 Quantifying the effect of population structure

Beyond individual risk-management, we found intrinsic differences in epidemic dynamics between populations that differed in their age or household size distributions. Comparing a “Florida-like” population to a “Texas-like” population (just “Florida” and “Texas”, hereafter), we find consistent, slight differences in the peak proportion of the population infectious at a given time (“Maximum Infectious”) and in the proportion of vulnerable individuals in the network that are infected over the course of the epidemic (fig. 2). Note that while vulnerable individuals make up a larger proportion of the population, on average, in Florida, we see a higher proportion of vulnerable individuals getting infected in the Texas population. This is a result of the higher rate of spread (also indicated by the higher peak proportion infectious) in the latter population, due in part to the larger average household size, and consequent higher number of strong within-household interactions.

**Figure 2:**
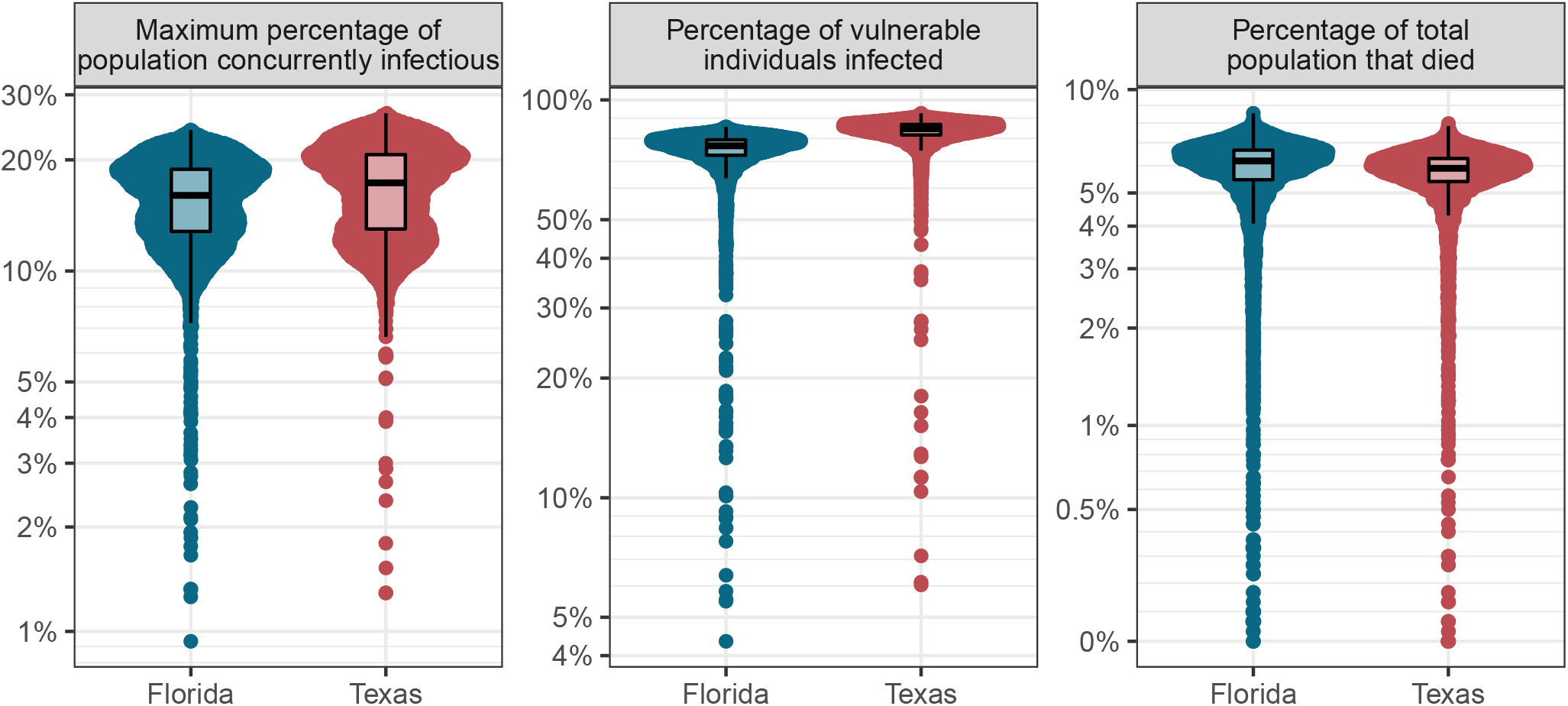
Comparing the difference in peak proportion infectious, overall prevalence among vulnerable individuals, and overall mortality between simulations of epidemics in two possible population structures, as characterized by age- and household-size distributions (see Supplementary Information and table 1). Florida has an (on average) older age distribution than Texas, while Texas has (on average) larger households. Each point represents a single simulated epidemic, conducted across a range of classmate- and co-worker transmission rates. Only simulations with no difference in behaviour based on vulnerability and only outcomes from epidemics resulting in greater than 5% of the total population being infected are shown (5 778 simulations for Florida, 6 297 for Texas). *n.b*. each panel has independent vertical axes limits. See Supplementary Information (section S3 and figs. S3 and S6) for analogous figures under different background transmission rates.

It is noteworthy that these small differences in peak proportion of the population concurrently infectious and proportion of vulnerable individuals infected were insufficient to fully counter the greater intrinsic mortality risk of the Florida population. This could be due, in part to the counter-acting effects of age and household size distributions. In short, the household size distribution of Texas tends to lead to larger outbreaks, but the larger proportion of vulnerable individuals in Florida means that a similar number of individuals die despite fewer total people being infected.

These results point to the importance of understanding trade-offs and nuances in population structure when implementing public health interventions. For instance, when distributing effort to minimize lives-lost, one must consider both properties of individuals in the population (*e.g*. what proportion of the population is at increased vulnerability to adverse outcomes?) and properties of the social contact network that interconnects those individuals (*e.g*. what are the most likely infection pathways by which those vulnerable people can become infected?). The efficacy and cost efficiency of any public health efforts will depend on understanding these nuances and their interaction. For instance, it may be more effective to isolate young family members of vulnerable individuals than vulnerable individuals themselves, since the latter tend to be older and have fewer social interactions to begin with [27].

Of course, the social and economic consequences of any intervention (which may be related to the total number of interactions removed under intervention) must likewise be taken into consideration [46]. Critically, the effects of public policies have unequal effects across a population: school closure most negatively affects less-educated families [47], wealthier individuals are more able to tolerate (and comply with) travel restrictions [48, 46], and already marginalized communities often bear the brunt of adverse medical outcomes [49]. Likewise, the distribution of underlying medical conditions is not distributed uniformly across the population, often correlating with race and socio-economic status [50, 51]. Thus, an intervention focused on (families of) vulnerable individuals, will necessarily also have disparate social and financial impact on these already marginalized individuals. In short, while this study focuses on generic effects of contact network structure on disease spread, real-world applications must additionally consider the specifics of which individuals are affected by a policy decision.

### 3.3 Quantifying effects of interaction types

While it is difficult to disentangle the web of interactions that make up modern societies, we used linear models to investigate the effects of a given change in interaction strength in one layer on the rate or extent of disease spread in the population.

We found that a change in the co-worker transmission rate consistently resulted in a larger change in epidemic outcome than a similarly sized change in the classmate transmission rate (fig. 3). For example, an increase in co-worker transmission rate will have approximately twice the effect on peak proportion infectious, total death burden, and time to that peak than will a similar increase in classmate transmission. When considering the scenario of no change in behaviour based on vulnerability, this ratio climbs to approximately 3. Consistent with results in fig. 1, we saw smaller slopes (and reduced differences between the effects of different network layers)for total number of deaths when households containing vulnerable individuals limited exposure. Consistent with fig. 2, we saw that changes in transmission rates tended to have a larger effect (*i.e*. model coefficient magnitude) on epidemic size in Texas, and on mortality in Florida (driven mostly by workplace interactions).

**Figure 3:**
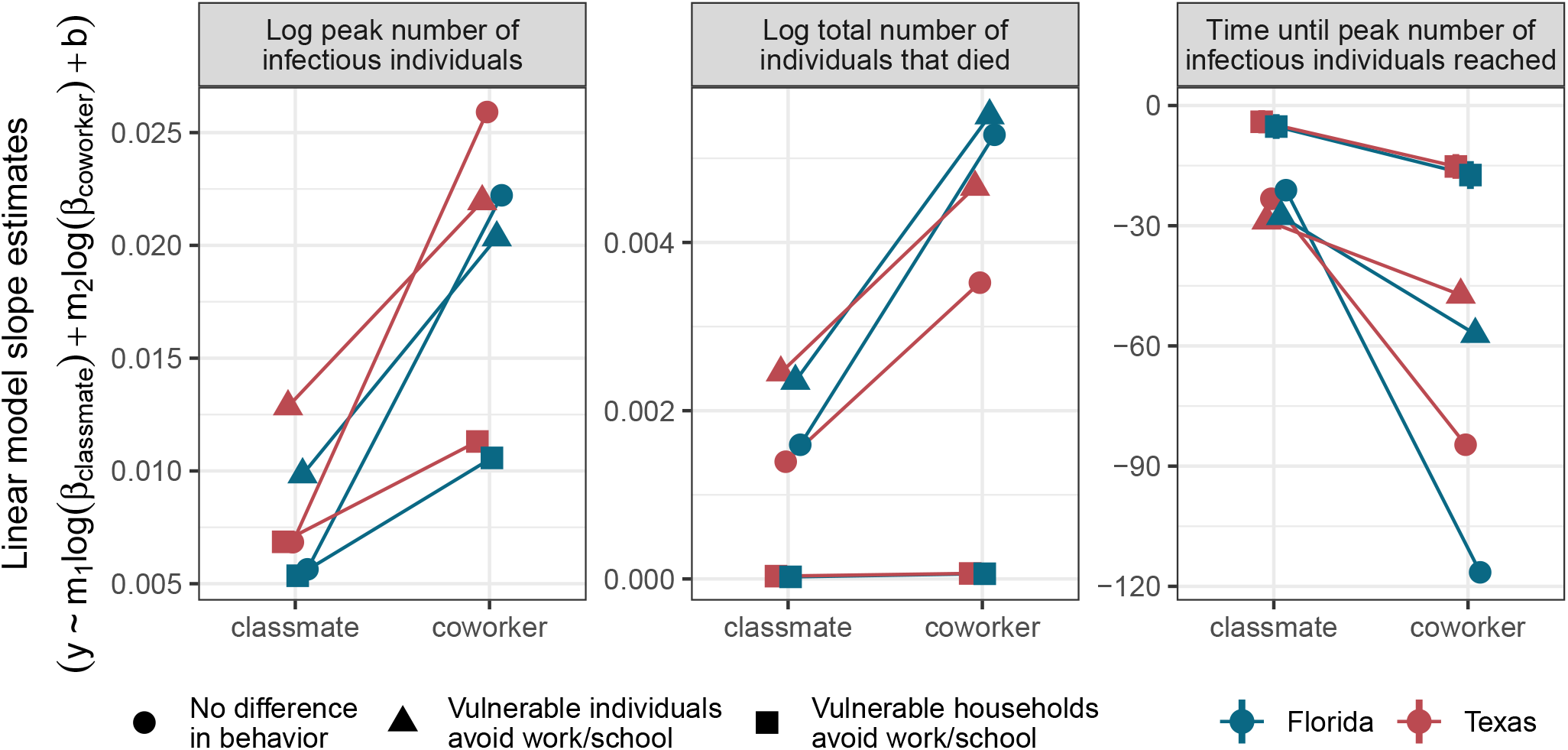
Quantifying the effect of changes in transmission rates on epidemic outcomes. The vertical axis indicates the value of the best-fitting coefficient for each transmission rate in a linear model of the form *y* ∼ *m*_1_*log*(*β*_*classmate*_)+*m*_2_*log*(*β*_*co* − *worker*_)+*b*, where *y* indicates an epidemic outcome measure, *m*_*n*_ is a fitted slope coefficient, *β*_*x*_ represents a transmission rate for interaction type *x*, and *b* is a fitted intercept coefficient. The horizontal axis distinguishes between the two coefficients (*m*_1_: “classmate”, or *m*_2_: “co-worker”). Facets distinguish between epidemic outcome measures, point shapes distinguish risk-tolerance regimes (*i.e*. rows in fig. 1), and point colours distinguish age and household size distribution locales (as in fig. 2). Vertical lines extending beyond the point extents indicate 95% confidence intervals for the slope estimates (most confidence intervals are obscured by the points). To ease interpretation, lines connect coefficient values across interaction types for results from models of the same risk-tolerance regime and locale. Points are slightly offset horizontally to reduce overlap. Note that a more positive slope in the left and center facets indicates a greater number of individuals infectious or died, respectively, while a more negative slope in the right facet indicates a faster rate of infection (less time to reach peak infectiousness). Only outcomes from epidemics resulting in greater than 5% of the total population being infected were included in the linear models (13 182 simulations for Florida, 14 844 for Texas). *n.b*. each panel has independent vertical axes limits. See Supplementary Information (section S3 and figs. S4 and S7) for analogous figures under different background transmission rates.

This is driven in part by the difference in the number of interactions in the network associated with each layer of the network. Because there are more individuals of working age than of school age, and because workplaces can potentially be much larger than classrooms, there tends to be more co-worker interactions in a given network than classmate interactions. While this is dependent upon the assumptions underlying construction of these simulated contact networks, it is also generically true of the real human contact networks that inspired our approach. In most real-world societies, there are more working-age individuals than children, and workplaces can potentially be orders of magnitude larger than school classrooms. The fact that these results seem to be robust to imbalance in interaction strength suggests that network structure (broadly construed) may have a larger role to play than pairwise interaction strength, at least for a highly transmissible disease like COVID-19.

While the surface-level implication of these results is that efforts should be focused on workplaces, rather than classrooms (and this is reinforced by evidence that, at least for early strains of SARS-CoV-2, transmission to, from, and among children may be less than that among adults; [35, 52, 53, 54]), schools still contribute meaningfully to disease spread, especially when considering some of the more recent (and more transmissible) strains of SARS-CoV-2 [55]. Importantly, these results are based on simulations where the internal networks for workplaces are very highly connected. Compartmentalizing workers, improving personal hygiene, ensuring adequate ventilation and air filtration, and supporting personal protective equipment usage can all alter the number and strength of interactions within the network. Such interventions would reduce the overall impact of workplace interactions on disease spread.

## 4 Limitations & future directions

While the networks used in this study are inspired by empirical human contact networks, there were nonetheless many assumptions built into their construction that are necessarily unrealistic. Future studies could, for example, consider differences in fine-scale network structure between interaction types, add additional explicit interaction layers, and increase network size to better reflect a whole urban area. In particular, one might nest individual classrooms within a less-strongly connected collection to represent a school, where interactions can likewise occur in public spaces such as the cafeteria or library [26, 56]. Such interactions are increasingly likely as schools relax their physical distancing requirements. Similarly, there might be differences in between- and within-classroom structure for differently aged students. Within workplaces, there might be a hierarchical network structure, where some peoples (*e.g*. managers) might interact with a collection of individuals that otherwise have little interaction with one another. Finally, we focused on only two types of interactions: those between classmates and those between co-workers. Clearly, there are myriad other ways in which individuals interact with one another, each of which might be structured in unique ways.

This study was in part limited by available data sources. While national-level data is readily available for most elements of our network generation, the same data for localities (even at the level of US states) is less accessible. An additional consideration is covariance between different aspects of population structure, where most data sources are segmented. For instance, one might assume that larger households have a lower average age (*i.e*. more children), yet age and household-size distributions are only available independently through the US Census American Community Survey.

In our disease model, we utilized disease parameters corresponding to the initial wave of COVID-19, despite substantial strain evolution since that time. Because our focus in this work is not on any one disease in particular, we opted to use an older strain for the increased availability and reliability of parameter estimates. These literature-based parameters additionally result in mortality being almost exclusively among vulnerable individuals—a trait we treated as binary and assigned based on post hoc empirical hospitalization rates. A more robust approach would be to consider the distribution of specific underlying health conditions within the population and their relative contribution to adverse outcomes. Critically, this approach also assumes constant mortality rates, disregarding a known relationship in which increased hospital occupancy results in worse disease outcomes [57].

## 5 A note on generality

In the midst of an ongoing pandemic of COVID-19, much of the inspiration for this work, and literature referenced herein has considered this one disease in particular. Nevertheless, the results presented here stem from a disease model that could be applied to many transmissible diseases with minimal modification. Even in the consideration of SARS-CoV-2, as new strains arise, resulting in altered rates of transmission, progression, recovery, and/or mortality, we expect the fundamental effects of network structure on disease spread to persist. For instance, as COVID-19 mortality rates have sequentially declined over the past two years, focus has shifted from mortality to hospitalization. As the drivers behind hospitalization and mortality are largely equivalent [58, 59], one could apply this same framework in the context of hospitalization. Likewise, vaccination (depending on efficacy) can be thought of as equivalent to either removing interactions (as in isolation) or reducing interaction strength (as in increased mask usage). Thus, omitting consideration of the additional benefits of reduced disease severity on the vaccinated individual, vaccination strategy could mirror consideration of physical distancing recommendations in this work. Importantly, when reduction of disease severity is additionally considered, previous work has suggested that prioritizing vulnerable individuals is most efficacious [60].

## 6 Conclusion

Our simulations reinforce the consequences of our highly connected, modern society on disease spread. In short, we find that decisions are rarely “personal” when it comes to public health, and the policies and health decisions of a population can have dramatic effects of the spread of disease. Action by vulnerable individuals in isolation does little to reduce their disease burden, suggesting that policies should additionally consider the potential for next-order transmission to vulnerable individuals from the less-vulnerable individuals that interact with them. Additionally, a population’s composition and social contact network structure can have marked effects on disease prevalence and mortality, though in our analysis these relationships were slight and sometimes resulted in counter-intuitive results whereby rapid disease spread can counteract the benefit of an otherwise less-vulnerable population. Finally, the structure of workplaces potentially provides greater avenues for disease spread than do schools, but these effects are highly dependent on both how workplaces/schools are structured, as well as the utilization and efficacy of non-pharmaceutical interventions in each of these contexts.

While over-interpretation of specific values should be avoided in purely simulation-based studies such as these, comparisons between different simulations can nevertheless provide insight into the relative importance of different components of a contact network on the rate and extent of disease spread. By comparing simulations across constrained axes of variation, such as types of interactions, differences in personal risk tolerance (or systemic structures and policies), and different population structures, we glean insights into how the different layers of social contact networks can have different levels of importance when it comes to containing epidemic spread. We can use this nuanced understanding to better inform and differentiate between public health strategies.

## Data Availability

All data produced in the present work are contained in the manuscript

## 7 Acknowledgements

We thank Janine Mistrick, Chris Wojan, Emily Schafsteck, Desiree Walton, and Lisa Meyer for feedback on early drafts of this work.

## Supplementary Information

### S1 Network data sources

To construct our plausible human contact networks, we relied on a range of publicly available data sources. For household sizes, we used state-specific data for Florida [S1] and Texas [S2], for the latter, we were only able to obtain accurate numbers for households with three or fewer occupants, so higher values were extrapolated from the overall average occupancy [S3]. Individual vulnerability was assigned based on age-specific COVID-19 induced mortality rates [S4]. To construct the underlying age distribution, we used United States American Community Survey data from 2019, specifically publications DP05: Demographic and Housing Estimates [S5]. Each individual in the network was probabilistically assigned into one of thirteen (less than 5 years old, 5-9, 10-14, 15-19, 20-24, 25-34, 35-44, 45-54, 55-59, 60-64, 65-74, 75-84, or greater than 84 years old) age classes based on state-specific age distributions. The distribution of ages across the network was modified from an initial random allocation to prevent the occurrence of households in which all individuals were children.

The school layer was constructed by taking all school-age individuals (5-9, 10-14, or 15-19 years old) and assigning them into age-class-specific classrooms of approximately 20 students per classroom [S6]. Within each classroom, networks were fully connected: all students had the same interaction strength with all other students in the classroom.

The workplace layer similarly considered all working-age adults (20-24, 25-34, 35-44, 45-54, 55-59, or 60-64 years old), subtracted a percentage (10%) based on unemployment rates in the spring-summer of 2020 [S7], and assigned the remainder to workplaces whose size was loosely based on the distribution of workplace sizes in the United States [S8]. This latter distribution was modified to remove especially small (less than 5 workers) and large (greater than 100 workers) work places. Within workplaces, as with classrooms, networks are fully connected: all workers have an equal level of contact with all other workers at the same workplace.

### S2 Locality network structure comparison

**Figure S1:**
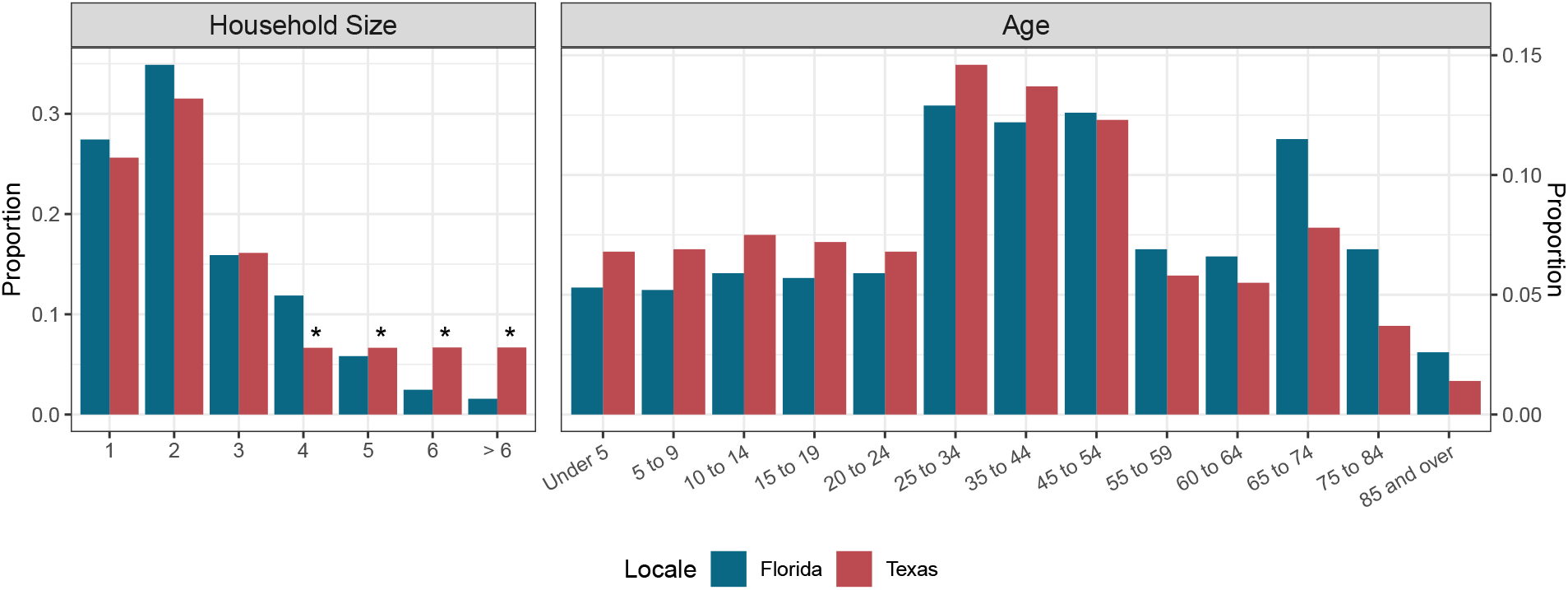
Household size (left) and age (right) distributions for each of the two locales used in the main text. *n.b*. left and right plots have different vertical axis limits. While Florida tends to have older citizens, Texas tends to have larger families (and consequently more within-household interactions; see also table 1). Asterisks denote values for Texas household size that were inferred to match an overall mean household size in the absence of precise data.

### S3 Alternative background transmission

#### S3.1 No background transmission (*β*_*background*_ = 0)

**Figure S2:**
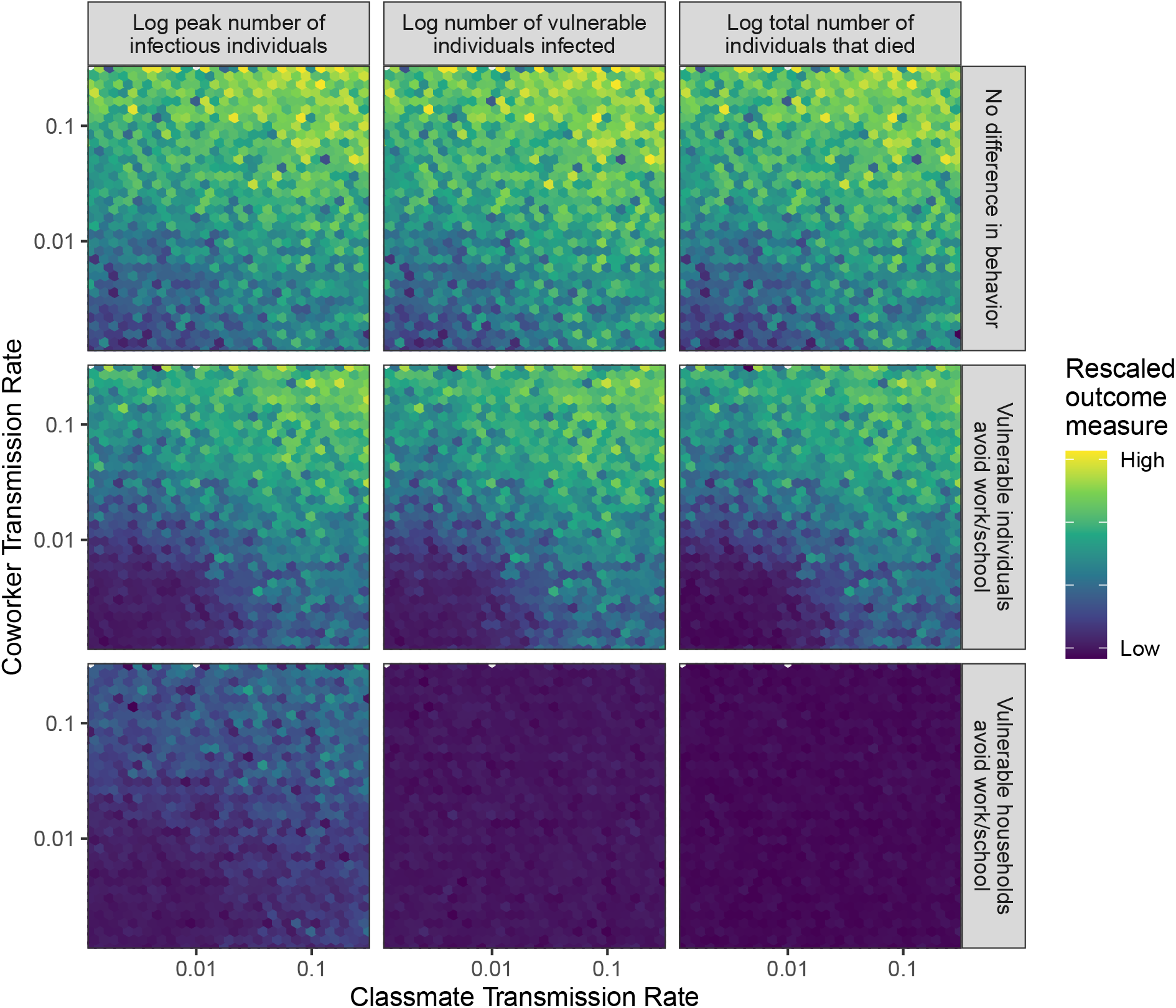
As fig. 1 in the main text, but with no background transmission. Relative epidemic outcome, quantified as the peak number of individuals infected (left), the number of vulnerable individuals infected (middle), or the total number of individuals that died (right) over the course of the simulation. Individuals in the network either: did not change behaviour in response to (contact with individuals with) vulnerability status (top), changed behaviour if they were vulnerable themselves (middle), or changed behaviour when a member of their household was vulnerable (bottom). Multiple points within each hexagon were averaged to produce the plotted value. Mean values were then log-scaled and normalized for each epidemic outcome such that the maximum value is 1 (yellow) and the minimum value is 0 (purple). Each panel consists of a heatmap showing the relative epidemic outcome of simulations spanning various levels of co-worker (vertical axis) and classmate (horizontal axis) transmission.

**Figure S3:**
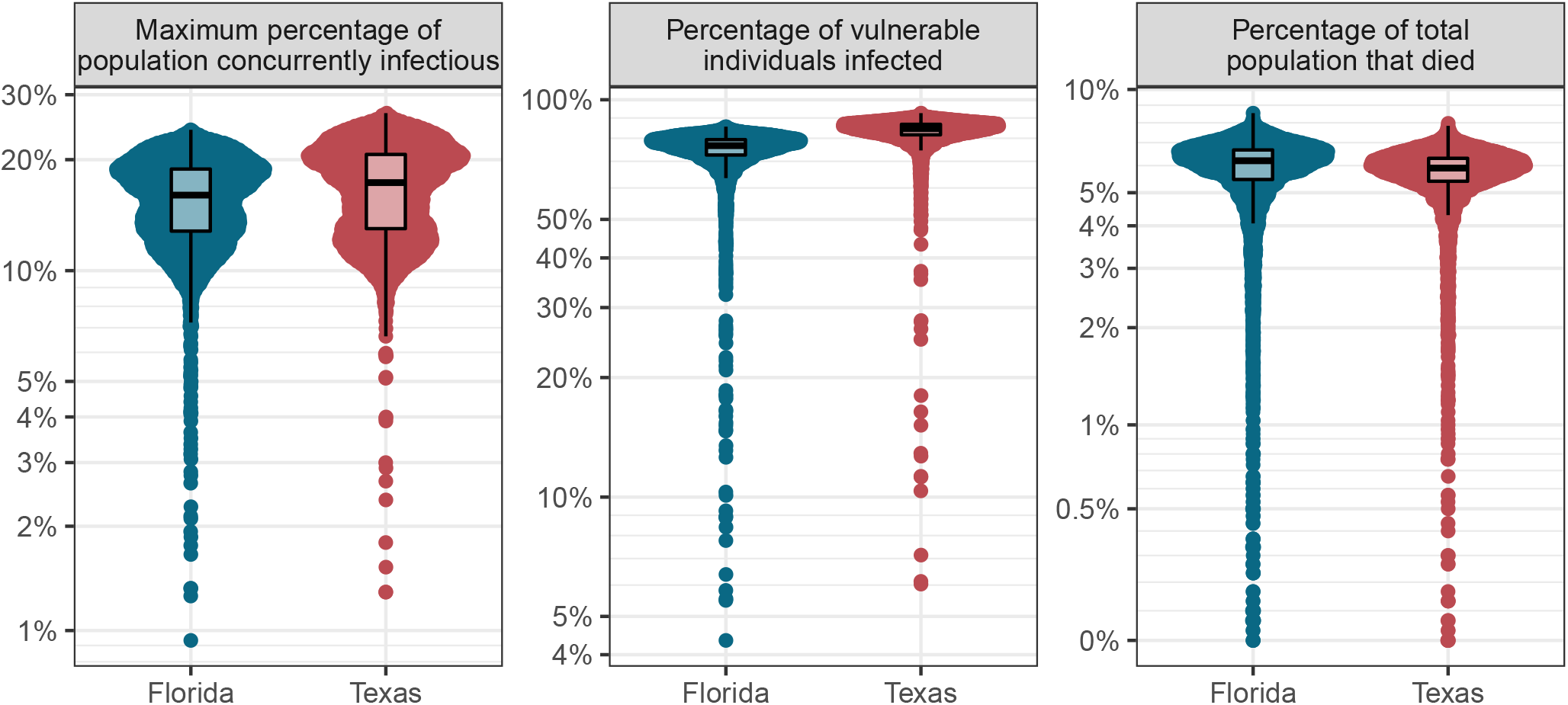
As fig. 2 in the main text, but with no background transmission. Comparing the difference in peak proportion infectious, overall prevalence among vulnerable individuals, and overall mortality between simulations of epidemics in two possible population structures, as characterized by age- and household-size distributions. Only simulations with no difference in behaviour based on vulnerability and only outcomes from epidemics resulting in greater than 5% of the total population being infected are shown.

**Figure S4:**
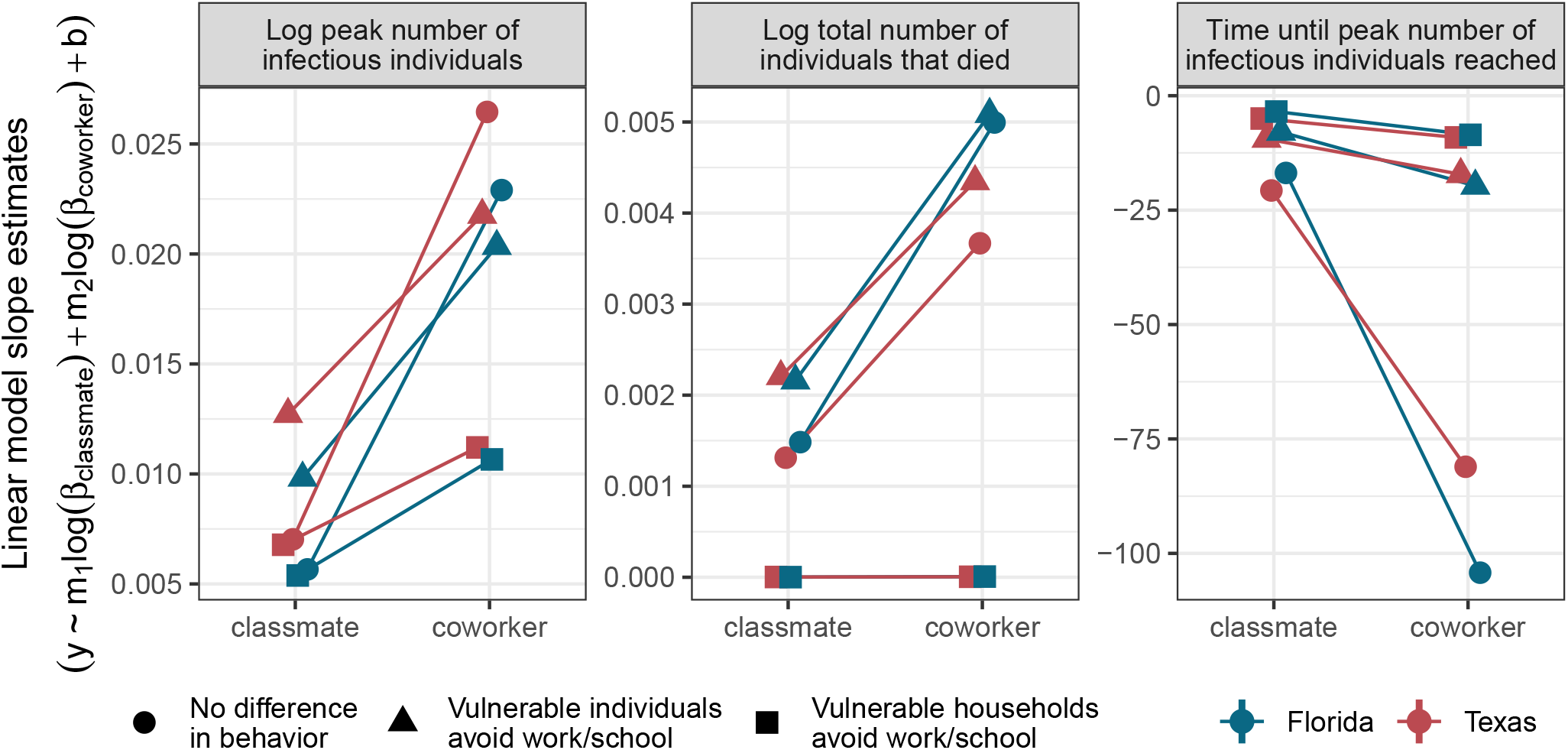
As fig. 3 in the main text, but with no background transmission. The vertical axis indicates the value of the best-fitting coefficient for each transmission rate in a linear model fit to simulation output. Facets distinguish between epidemic outcome measures, point shapes distinguish risk-tolerance regimes (*i.e*. rows in fig. 1), and point colours distinguish age and household size distribution locales (as in fig. 2). Vertical lines extending beyond the point extents indicate 95% confidence intervals for the slope estimates (some confidence intervals are obscured by the points). To ease interpretation, lines connect coefficient values across interaction types for results from models of the same risk-tolerance regime and locale. Points are slightly offset horizontally to reduce overlap. Only outcomes from epidemics resulting in greater than 5% of the total population being infected were included in the linear models.

#### S3.2 High background transmission (*β*_*background*_ = 0.1)

**Figure S5:**
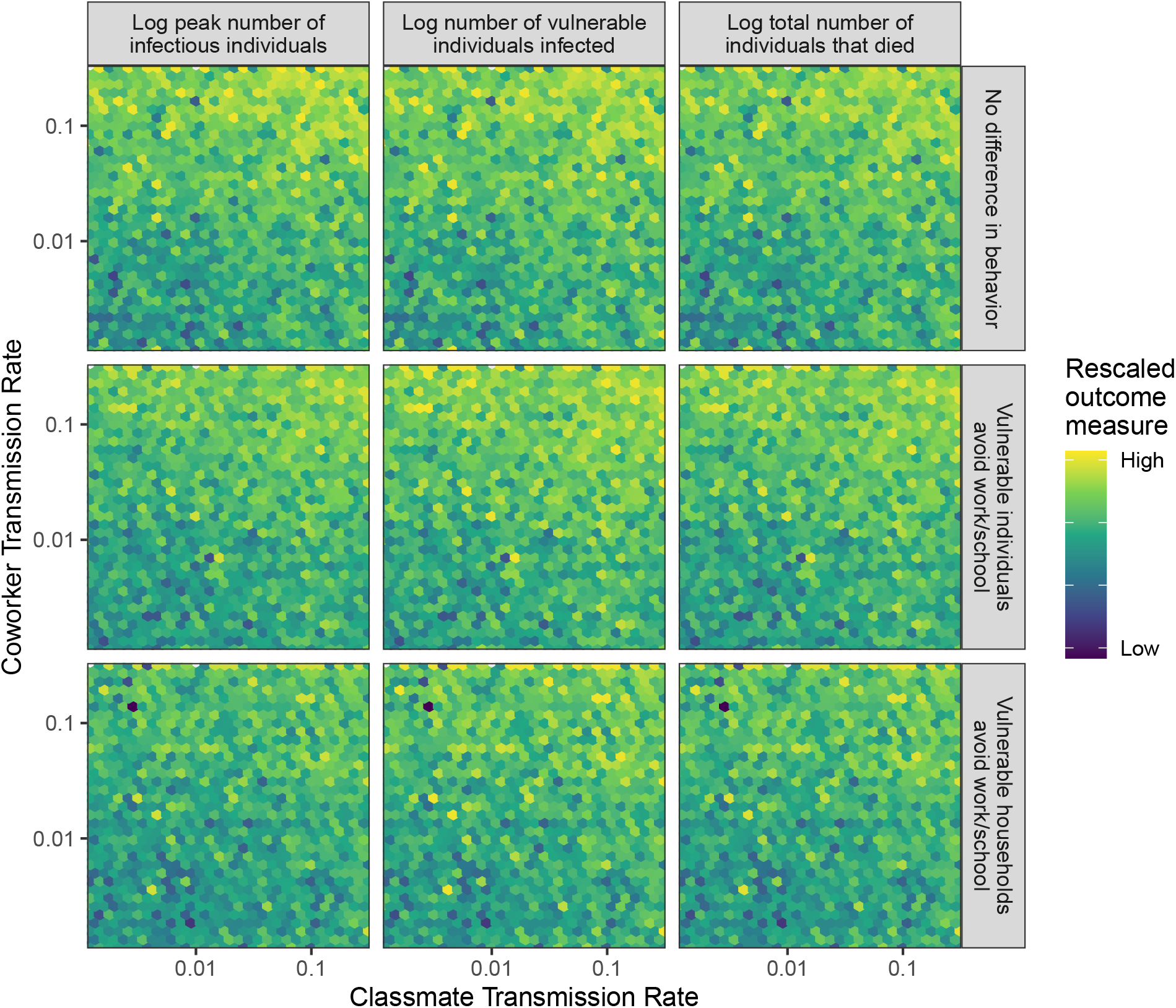
As figs. 1 and S2, but with background transmission set to *β*_*background*_ = 0.1*/N*.

**Figure S6:**
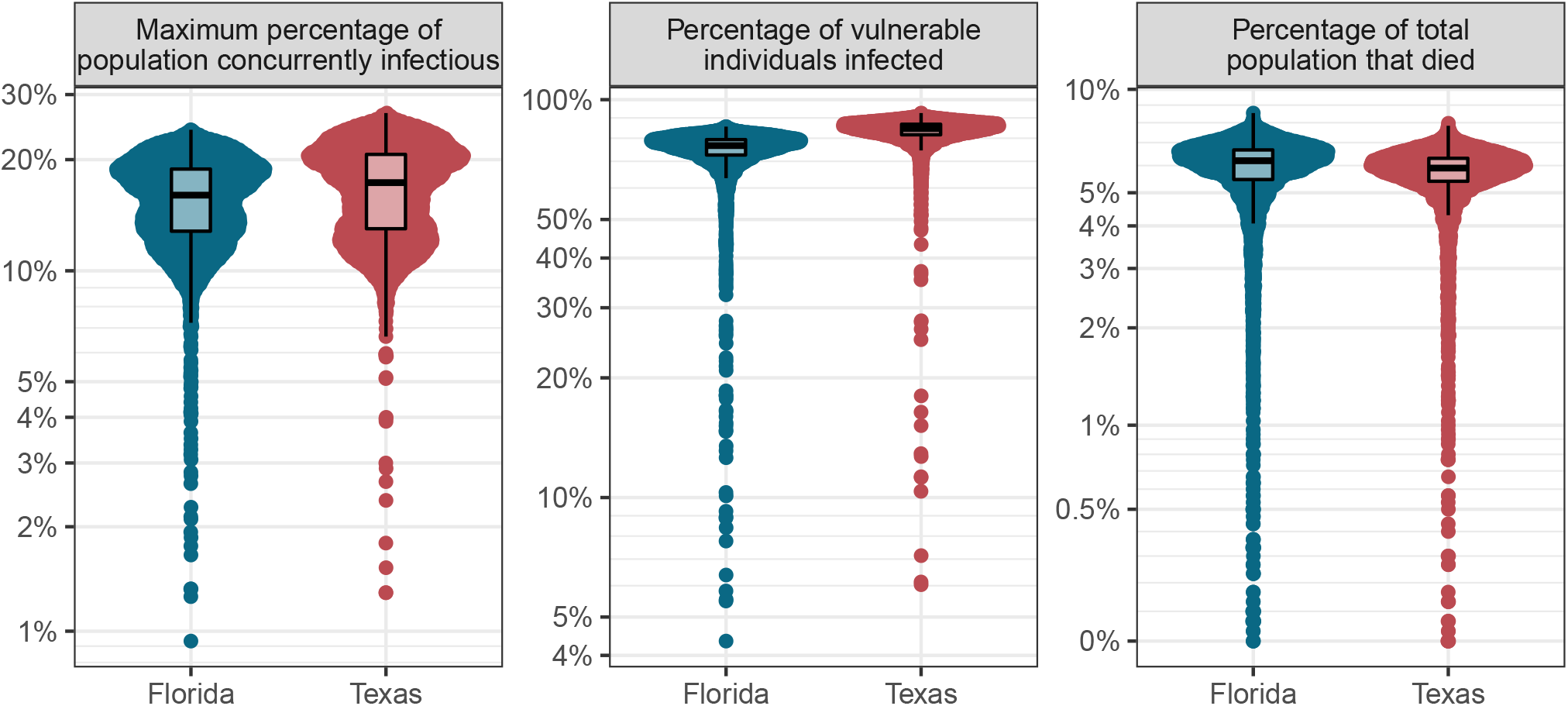
As figs. 2 and S3, but with background transmission set to *β*_*background*_ = 0.1*/N*.

**Figure S7:**
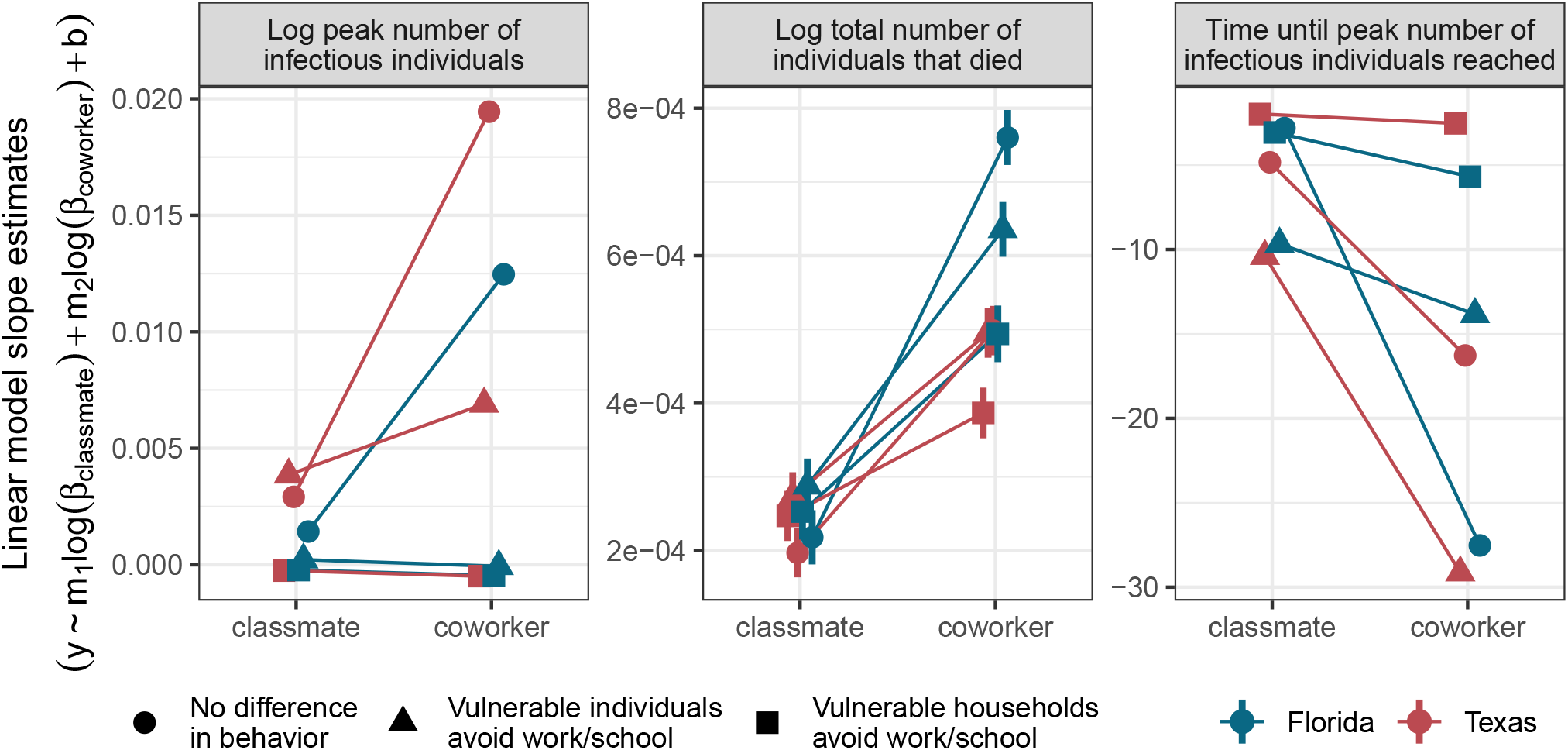
As figs. 3 and S4, but with background transmission set to *β*_*background*_ = 0.1*/N*.

### S4 Simulation code accessibility details

All simulations were conducted in C++ version 8.1.0, with data manipulation and plotting done in R version 4.2.0 [S9], with the use of R packages: assertthat [S10], ggbeeswarm [S11], kableExtra [S12], patchwork [S13], Rcpp [S14, S15, S16], tidygraph [S17], tidyverse [S18], and scales [S19].

An application for visualizing our synthetic community network structure and simulating disease spread (including the manipulation of disease parameters) is available online: https://z.umn.edu/LINCS [S20]. Code to replicate all aspects of these analyses is available online: https://github.com/mjsmith037/Layered_Interactions_COVID_Model.

### S5 Link removal irrespective of (proximate) vulnerability

**Figure S8:**
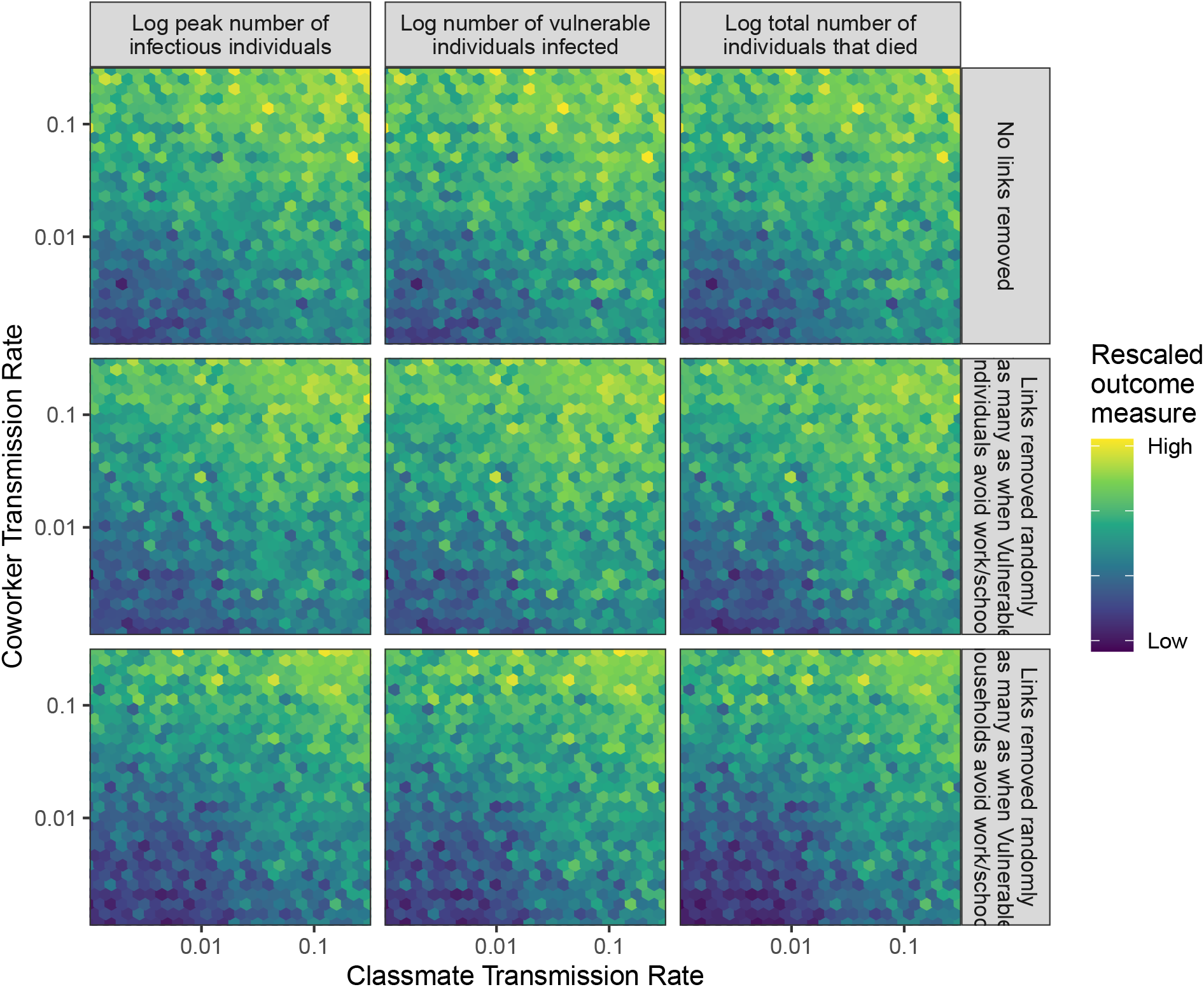
As fig. 1 in the main text, but rather than removing links in association with an individual’s contact with vulnerable individuals, the same number of links were removed randomly from the same interaction types as in fig. 1.

